# Peculiar morphologic characteristics of adrenal veins and similarity to fibromuscular dysplasia of artery: implication of pathogenesis

**DOI:** 10.1101/2023.01.04.23284197

**Authors:** Peilin Zhang, Daniel Rafii, Minerva Romero Arenas

**Author notes:** Correspondence: Peilin Zhang, MD, Ph.D., Pathology, NYP- Brooklyn Methodist Hospital, Brooklyn, NY 11215, Weill Cornell Medicine. **Funding:** No funding is received for this study.

## Abstract

**Background:** Fibromuscular dysplasia (FMD) is a peculiar abnormality of arterial wall with characteristic bead-like features on angiogram that commonly occurs in renal arteries and carotid arteries. The morphologic features of FMD of arteries share significant similarities to those described for adrenal veins. The pathogenic mechanism of FMD remains largely unknown and genetic susceptibility appears important.

**Methods:** We examined the morphologic characteristics of adrenal vessels to compare with fibromuscular dysplasia and segmental arterial mediolysis. We have retrospectively reviewed 30 cases of adrenalectomy specimens associated with or without adrenal neoplasms including cortical adenomas and pheochromocytomas regarding the histomorphologic features of adrenal vessels within the normal tissue and the tumors.

**Results:** Adrenal veins showed characteristically asymmetrical muscle bundles in normal adrenal glands in all cases except for one adrenal cortical adenoma in which normal adrenal gland was absent. These morphologic features shared significant similarities with those of FMD of arteries. A spectrum of vascular changes including hyalinization, intimal fibroplasia, myxoid degeneration and atherosis was observed in 10 of the 15 cases of adrenal cortical adenomas and 5 of the 10 cases of pheochromocytomas. Most common complications were hemorrhage/hematoma and thrombosis. There is no statistically significant differences in patients’ baseline characteristics including race/ethnicity, marital status, blood pressure and body mass index (BMI).

**Conclusion:** The unique vascular changes in adrenal glands and adrenal neoplasms suggests the importance of tissue specific milieu likely related to adrenal hormones/hormone receptors, providing direction of further investigation of pathogenic mechanisms and potentially management of FMD and similar diseases.

## Introduction

Human adrenal central vein is located deep in the medulla characterized by unique longitudinal asymmetrical smooth muscle bundles developed over age [1-4]. There is no circular smooth muscle bundle within the adrenal central vein. The significance of this peculiar morphologic feature is unknown and it was hypothesized that asymmetrical longitudinal muscular bundles controls epinephrine release and hypertrophy of the muscle over time is protective to the hypertensive patients [1, 2, 5]. Fibromuscular dysplasia (FMD) is a peculiar abnormality of arterial wall characterized by small dissecting aneurysms with diagnostic bead-like features on imaging (angiogram) [6, 7]. Pathogenesis of FMD remains elusive but genetic susceptibility appears to play an important role [8]. FMD can occur at any anatomic site and it most commonly occurs in renal or carotid arteries with variable severity of arterial stenosis/ectasia/aneurysms [9-12]. Histologically, FMD shows variable thickness of arterial wall due to deficient muscular media or/and adventitia with non-inflammatory non-atherosclerotic changes, and these morphologic changes include fibroblastic proliferation (fibroplasia) or focal smooth muscle destruction/hyperplasia [13-15]. Histopathological and radiologic correlation of FMD had been elegantly demonstrated, and FMD is a radiographic morphologic diagnosis today [7, 15]. Pathological examination for clinical diagnosis of FMD is not required, although FMD itself appears a histologic term. Segmental arterial mediolysis (SAM), another similar vascular abnormality with similar morphologic and clinical features, is believed to be a precursor lesion of FMD [16, 17]. SAM is also a histologic term, and it has been described in many anatomic sites, but the majority of the literature was case report with various clinical complications and outcomes [18-21]. Pathogenesis of SAM appears related to vasospasm and adrenal hormones/adrenergic receptor distribution within the target tissue [22, 23].

We have examined a spectrum of FMD/SAM lesions of adrenal vessels including normal adrenal veins within the adult adrenal gland tissue associated with or without neoplasms including adrenal cortical adenomas or pheochromocytomas. We showed that there is significant morphologic similarities between the normal adrenal veins and FMD of arteries. A spectrum of vascular changes within the adrenal cortical adenomas and pheochromocytomas may pathogenically relate to the hormones secreted by the tumor cells. Our findings appear to support the unification of FMD and SAM into a spectrum of a single entity and suggest similar pathogenic pathway related to abnormalities of adrenal hormone actions and/or hormonal receptors.

## Materials and Methods

The study was approved by the Institutional Review Board (IRB) at New York Presbyterian Brooklyn Methodist Hospital (1592673-1). All adrenalectomy specimens from 2018 to September 2022 for neoplastic or non-neoplastic lesions were included in the study. Routine paraffin-embedded tissues and Hematoxylin & Eosin (H&E) stained slides were examined by light microscopy. The patient’s original diagnoses was retrieved and recorded in the Excel spreadsheet (Microsoft Corporation). The presence of the spectrum of vascular changes was also recorded in Excel spreadsheet (Microsoft Corporation) and patients’ clinical data including age, sex, race/ethnicity, marital status and body mass index were subsequently retrieved from the medical records. Maternal racial data were retrieved from the medical record according to the US census criteria as Asians, non-Hispanic Black, Hispanic, and non-Hispanic white. Our racial data also included “others”, “unknown”, or “declined” as one group. Marital status was listed as married, single, divorced, or others including those of unknowns and declined to respond. Statistical analysis was performed by using baseline characteristic table in R-package (http://statistics4everyone.blogspot.com/2018/01/.html).

## Results

Adrenal veins with characteristic asymmetrical longitudinal smooth muscle bundles were identified initially as incidental findings in normal adult adrenal tissues in the region between the adrenal cortical tissue and medullary tissue associated with adrenal cortical adenomas/pheochromocytomas during routine pathology examinations. These morphologic changes are characterized by gapping muscular media with nodular smooth muscle hyperplasia [1, 2]. In some areas, the smooth muscle media layer was diminished in half of the luminal wall with thin layers of endothelial cells (Figure 1). The elastic layers of adrenal arteries were diminished or absent (Figure 2), and some areas of the elastic tissue appear to be totally disorganized and undiscernible. The expression of smooth muscle myosin heavy chain (SMYOHC) by immunohistochemistry in muscular media was uneven and disarrayed with large areas of gapping of arterial wall and intact endothelial cell lumen positive for CD31 by immunohistochemistry (Figure 2). These morphologic features were consistent with classic description of normal adrenal central veins [1, 2]. These morphologic features are also indistinguishable from those described in fibromuscular dysplasia of renal or carotid arteries [14, 15]. These morphologic features were only observed inside adrenal gland tissue, but not in arteries in the soft tissue surrounding adrenal glands.

**Figure 1:**
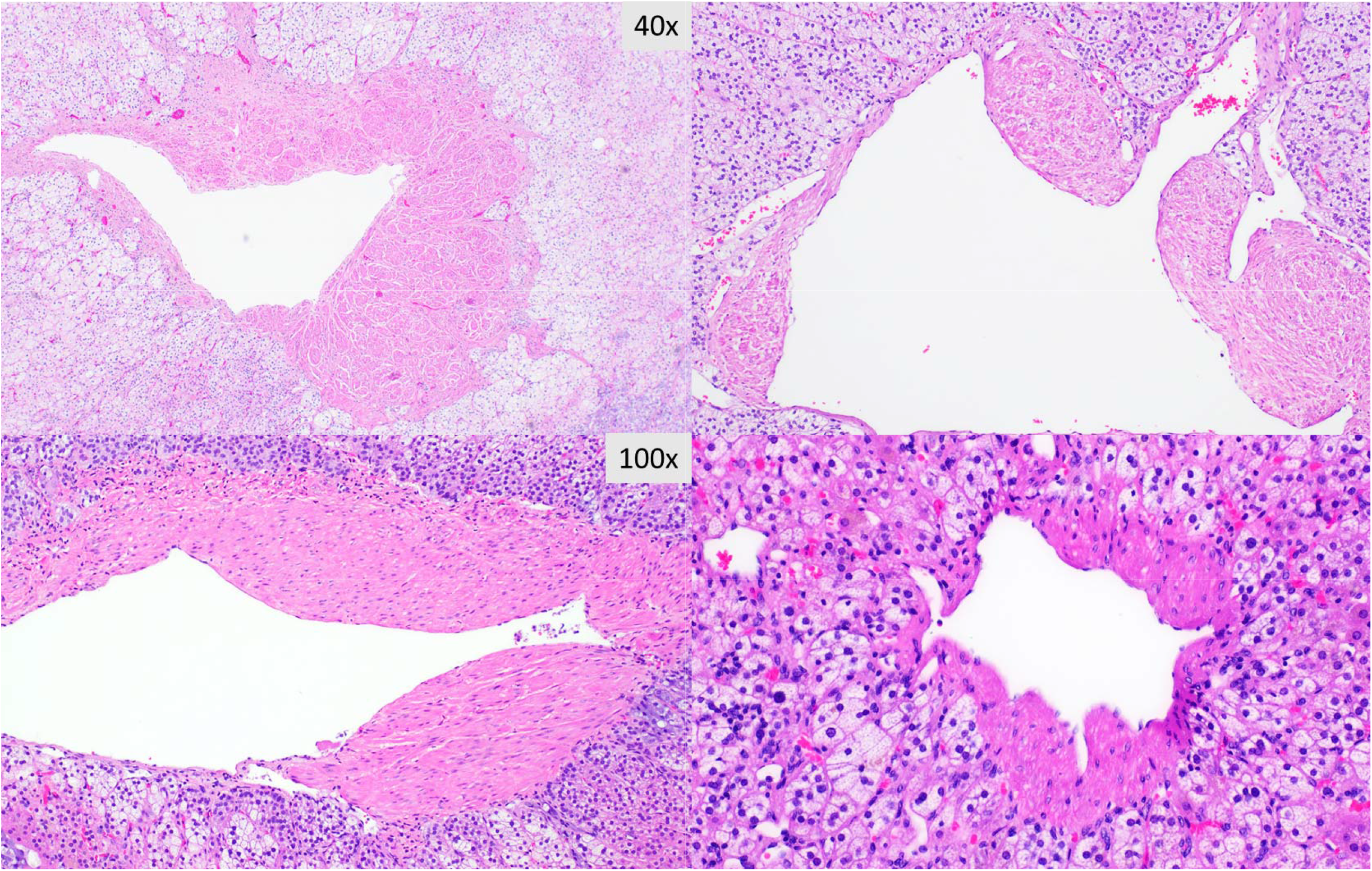
Morphologic features of adrenal veins with asymmetrical smooth muscle bundles within normal adrenal glands at various magnifications.

**Figure 2:**
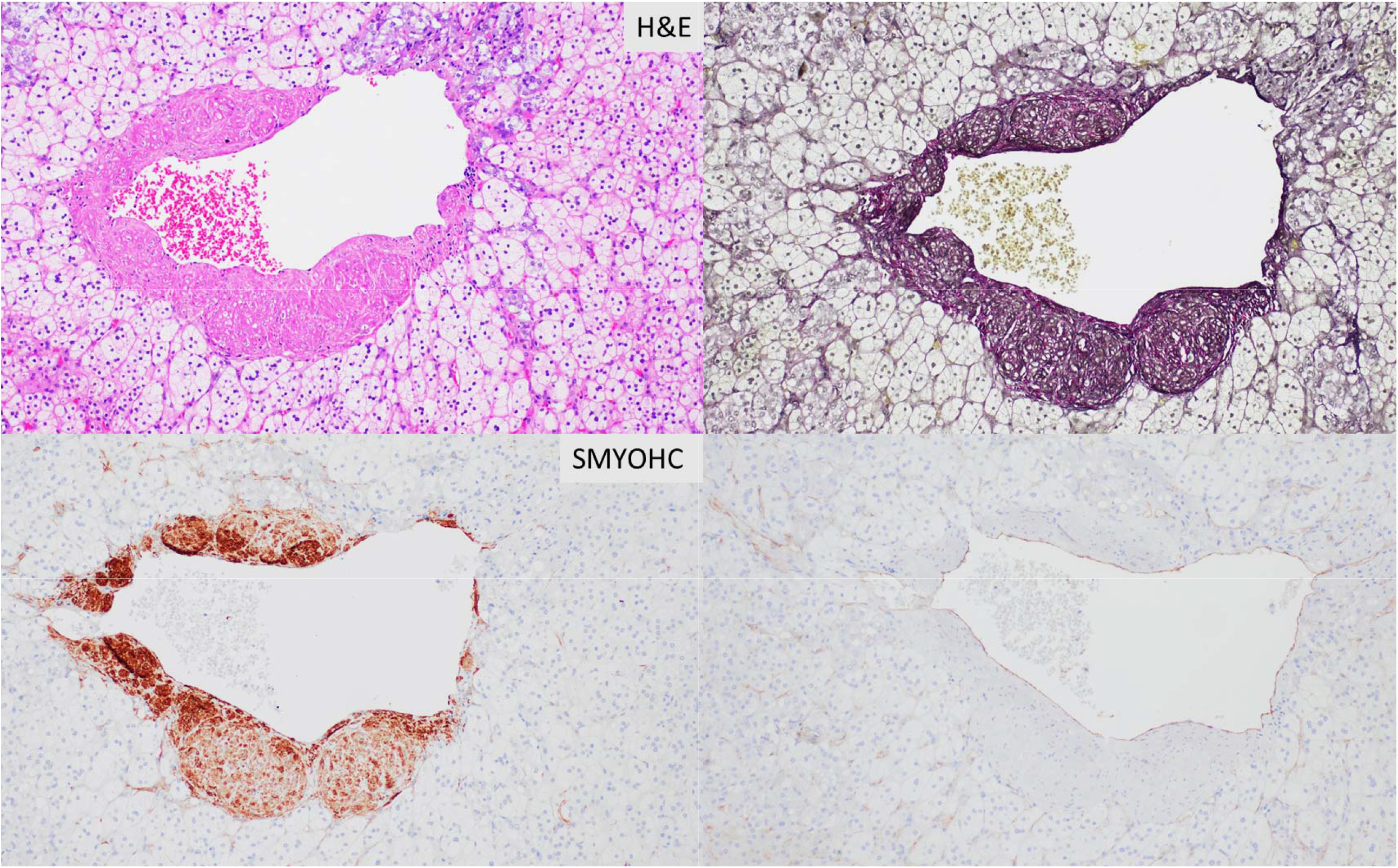
Adrenal veins with hematoxylin and eosin stain (H&E), elastin stain (Verhoeff’s Van Gieson /EVG), SMYOHC and CD31 stains by immunohistochemistry. All magnification 200x.

In adrenal cortical adenomas, the muscular media of the arteries was hyalinized with foamy macrophages in the lumen and wall, and these macrophages were positive for CD68 by immunohistochemistry (Figure 3). In some areas intimal fibroplasia, myxoid and fibrinoid medial degenerations/necrosis were noted (Figure 4), and these features were mostly observed in adrenal cortical adenomas. These morphologic characteristics were consistent with segmental arterial mediolysis (SAM) which were also similar to acute atherosis of macrophage type of placental membranes described in preeclampsia [22, 24, 25]. In pheochromocytomas, the arteries were mostly thin walled with significant reduction of muscular wall thickness and areas of “blood lakes” without discernable luminal endothelial linings. Various degrees of myxoid or fibrinoid degeneration of muscular media and fibroplasia of the intimal layer were common in pheochromocytomas, and normal muscular arteries were infrequent in either cortical adenomas or pheochromocytomas (Figure 4).

**Figure 3:**
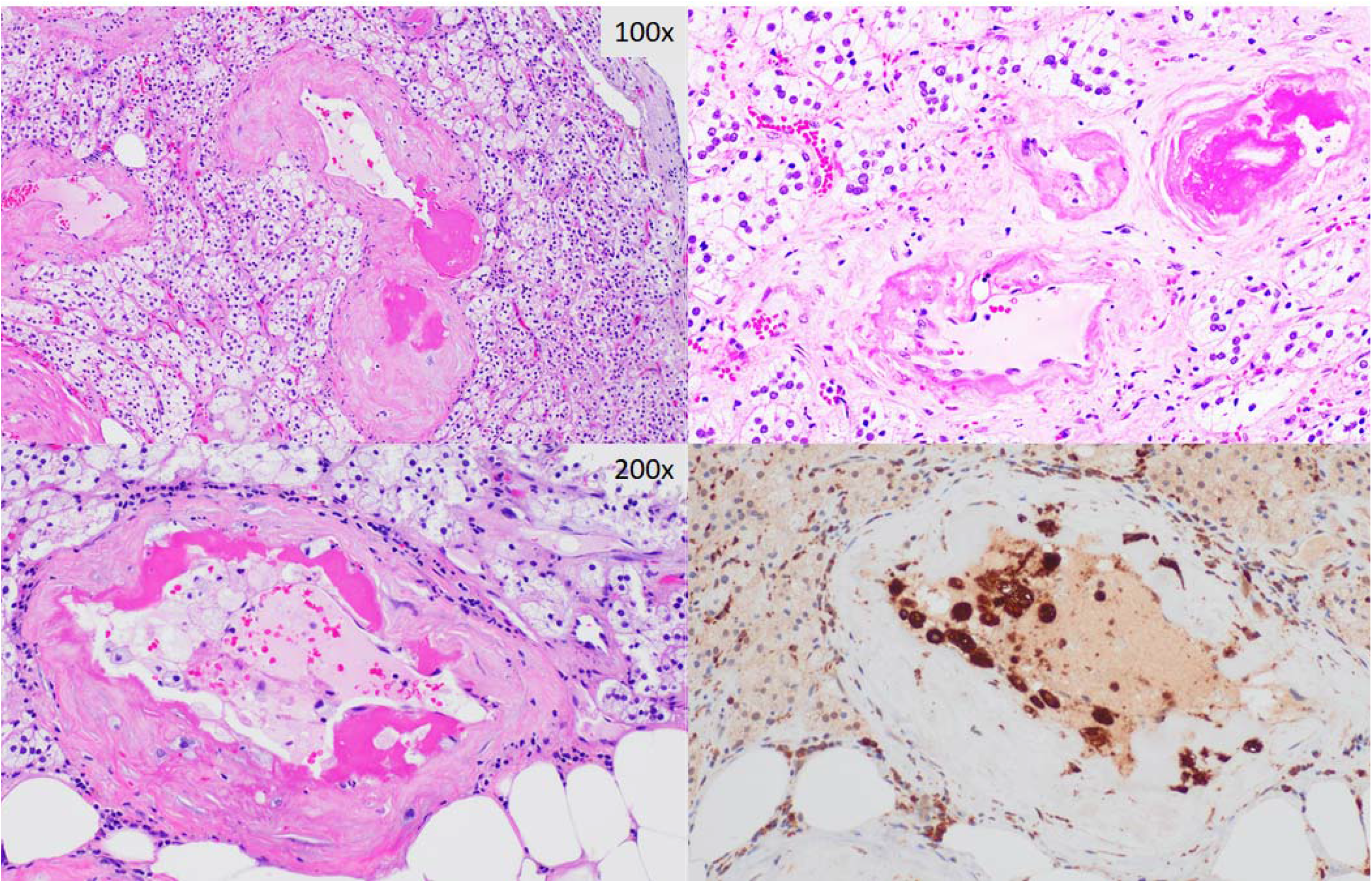
Variation of SAM and FMD with hyalinization of arterial walls with intraluminal or intramural macrophages and CD68 immunohistochemical stain.

**Figure 4:**
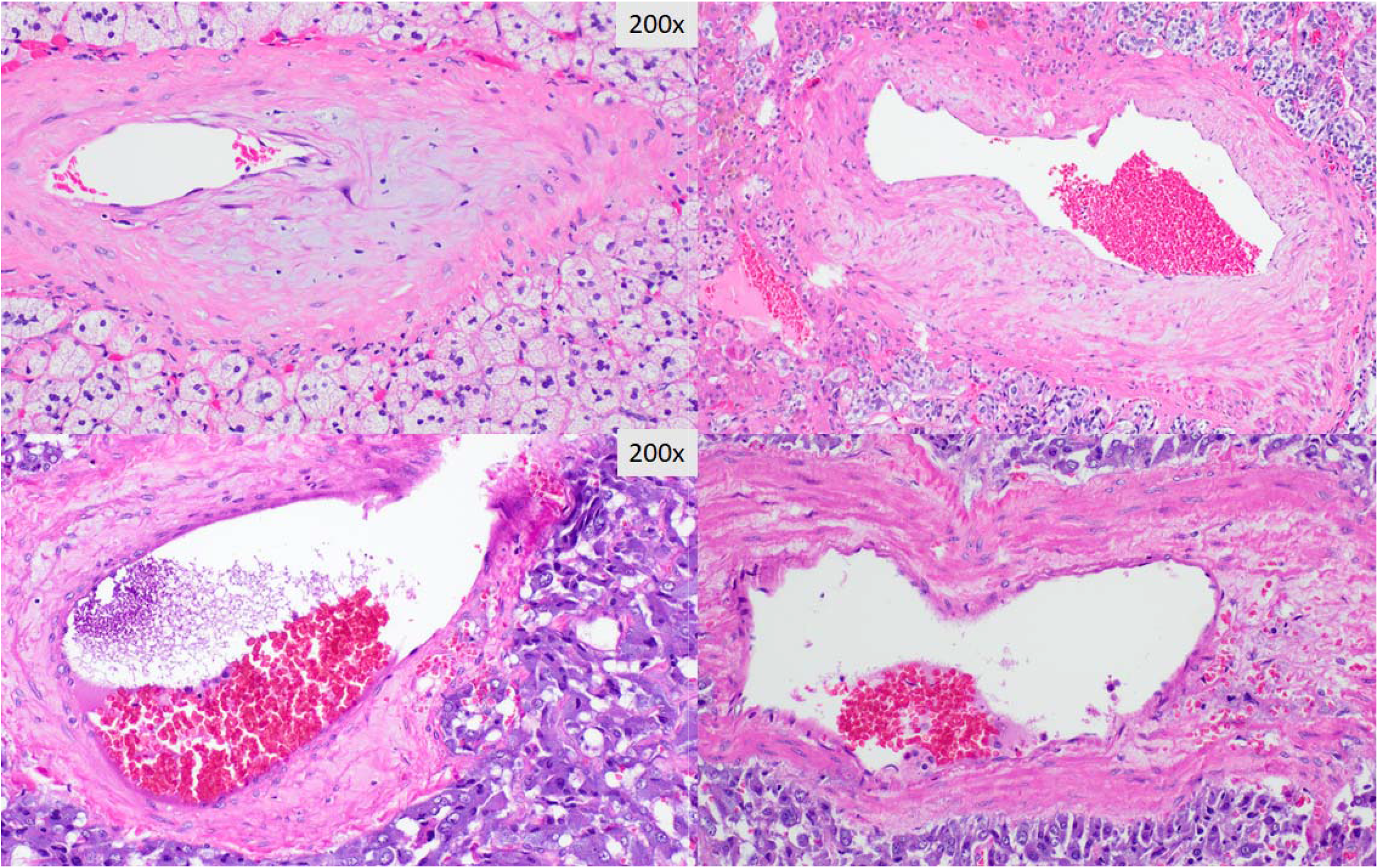
Intimal fibroplasia and myxoid degeneration of muscular media (SAM/FMD) of adrenal vascular walls in pheochromocytomas. All magnification 200x.

A total 30 cases of adrenalectomy specimens were reviewed retrospectively, and the patients’ baseline characteristics were shown in Table 1. All normal adult adrenal tissues associated with or without adrenal tumors including cortical adenomas and pheochromocytomas showed asymmetrical muscle bundles of the adrenal veins. Asymmetrical muscle bundles can be occasionally identified but not common in tumor tissues in either adrenal cortical adenoma or pheochromocytoma. However, SAM was commonly observed in most of adrenal cortical adenomas and pheochromocytomas. The most common complications in our cases were hemorrhage and vascular thrombosis (Table 1). There were no FMD or FMD-like lesions in the arteries outside adrenal gland in all cases with or without adrenal neoplasms. Review of fetal autopsy cases showed normal adrenal vessels in fetuses, and no FMD or FMD-like lesions or asymmetrical muscle bundles were identified in 9 fetuses of 15 to 37 gestational weeks.

**Table 1:** Characteristics of adrenal tumors and vascular pathology.

| Adrenal tumors | Adenoma<br>(N=15) | Cyst/hematoma<br>(N=5) | Pheochromocytoma<br>(N=10) | Total<br>(N=30) | p value |
| --- | --- | --- | --- | --- | --- |
| Race/Ethnicity |  |  |  |  | 0.41 |
| - Asian | 2 ( 13%) | 0 ( 0.0%) | 2 ( 20%) | 4 ( 13%) |  |
| - Black | 5 ( 33%) | 2 ( 40%) | 6 ( 60%) | 13 ( 43%) |  |
| - Hispanic | 2 ( 13%) | 2 ( 40%) | 1 ( 10%) | 5 ( 17%) |  |
| - White | 6 ( 40%) | 1 ( 20%) | 1 ( 10%) | 8 ( 27%) |  |
| Marital status |  |  |  |  | 0.09 |
| - Divorced | 0 ( 0.0%) | 1 ( 20%) | 0 ( 0.0%) | 1 ( 3%) |  |
| - Married | 4 ( 27%) | 2 ( 40%) | 6 ( 60%) | 12 ( 40%) |  |
| - Single | 11 ( 73%) | 2 ( 40%) | 3 ( 30%) | 16 ( 53%) |  |
| - Unknown | 0 ( 0.0%) | 0 ( 0.0%) | 1 ( 10%) | 1 ( 3%) |  |
| Age (year) | 63 [ 54; 65] | 61 [ 60; 69] | 64 [ 48; 69] | 62 [ 53; 68] | 0.68 |
| Sex |  |  |  |  | 0.57 |
| - Female | 8 ( 53%) | 4 ( 80%) | 6 ( 60%) | 18 ( 60%) |  |
| - Male | 7 ( 47%) | 1 ( 20%) | 4 ( 40%) | 12 ( 40%) |  |
| Body mass index (BMI) | 35 ± 7 | 31 ± 5 | 29 ± 7 | 33 ± 7 | 0.16 |
| Blood pressure (Systolic) | 144 ± 26 | 143 ± 13 | 128 ± 17 | 138 ± 22 | 0.20 |
| Blood pressure (Diastolic) | 82 [76; 92] | 86 [82; 90] | 76 [74; 77] | 80 [76; 88] | **0.05** |
| Mean arterial pressure | 98 [92; 114] | 104 [100; 111] | 92 [88; 96] | 96 [92; 108] | 0.09 |
| **Segmental mediolysis (SAM)** |  |  |  |  |  |
| - Atherosclerosis | 5 ( 33%) | 1 ( 20%) | 0 ( 0.0%) | 6 ( 20%) | 0.13 |
| - Hyalinization | 8 ( 53%) | 0 ( 0.0%) | 0 ( 0.0%) | 8 ( 27%) | **<0.01** |
| - Intimal fibroplasia | 1 ( 7%) | 0 ( 0.0%) | 2 ( 20%) | 3 ( 10%) |  |
| - Myxoid degeneration | 5 ( 33%) | 0 ( 0.0%) | 4 ( 40%) | 9 ( 30%) | 0.26 |
| Total SAM | 10 ( 67%) | 1 ( 20%) | 5 ( 50%) | 16 ( 53%) | 0.19 |
| **Fibromuscular dysplasia (FMD)** | 14 ( 93%) | 5 ( 100%) | 10 ( 100%) | 29 ( 97%) | 0.60 |
| Hemorrhage | 6 ( 40%) | 3 ( 60%) | 5 ( 50%) | 14 ( 47%) | 0.72 |
| Thrombosis | 6 ( 40%) | 2 ( 40%) | 3 ( 30%) | 11 ( 37%) | 0.87 |
| SAM in tumor | 11 ( 73%) | N/A | 6 ( 60%) | 18 ( 60%) | 0.11 |
| Muscular artery in tumor | 1 ( 11%) | N/A | 5 ( 50%) | 6 ( 27%) | 0.09 |

## Discussion

We have examined the adrenal vessels of all cases and the morphologic features of adrenal veins were similar to those described historically [2-4]. These peculiar morphologic features of adrenal veins are developmentally regulated over age and it has been postulated to be protective in releasing epinephrine in hypertension [1, 2]. More importantly these peculiar morphologic features of adrenal veins were similar to those of FMD lesions of arteries described in other anatomic sites such as renal or carotid arteries [7]. FMD is a radiographic morphologic diagnosis today with functional significance, and the etiology and pathogenesis remain elusive to date [7, 13, 15]. In adrenal cortical adenomas, the arterial walls were mostly hyalinized with occasional intraluminal or intramural macrophages that were positive for CD68 by immunohistochemistry. The features of hyalinized arterial walls with macrophages were classic and diagnostic of acute atherosis of macrophage type in placental membranes in preeclampsia [24, 25]. Acute atherosis was first described in the fetal membranes of placentas from preeclampsia, and the etiology of acute atherosis remains elusive although extensive studies has been conducted in the last half century [26-29]. In pheochromocytomas, the arterial walls were significantly diminished in thickness (thin-wall) with occasional myxoid/fibrinoid degeneration of muscular media and intimal fibroplasia. Normal muscular arteries were uncommon in either cortical adenomas or pheochromocytomas. These morphologic features of FMD and SAM were also similar to those described for Moyamoya disease or Moyamoya syndrome of the brain [30-33]. Adrenal gland is unique that adrenal hormones are secreted including adrenalins (epinephrine and norepinephrine), and these hormones were shown to be important for development of SAM in animal models [23, 34]. Adrenal gland also secrets corticosteroids, and corticosteroids were critical in development of normal adrenal medulla in genetic mice models [35, 36]. The relationship between corticosteroids and vascular smooth muscle integrity is not clear clinically, although molecular studies have been extensive [37-39].

Clinically, there are case reports of FMD associated with pheochromocytomas but to our knowledge there were no clinical association between adrenal cortical adenoma with FMD and no histopathological analysis of adrenal vessels in cortical adenomas or pheochromocytomas were reported to date [40, 41]. Our data of adrenal vessels in normal adrenal tissue and adrenal tumors suggest the local milieu within adrenal glands or adrenal neoplasms including cortical adenomas or pheochromocytomas may play important roles in shaping the arterial walls and maintenance of arterial function, and the hormones produced by adrenal glands may have deleterious effects on arterial smooth muscle integrity. Alternatively, hormonal actions are related to hormone receptor functions and distributions, and variations of adrenal hormone receptors in arterial smooth muscle media may play roles in vascular functions and pathogenesis of FMD at various anatomic sites [42]. The similar morphologic features of adrenal vein, FMD, SAM, atherosis of preeclampsia and Moyamoya disease/syndrome may suggest similar underlying pathogenic mechanisms related to adrenal hormone actions or hormone receptor signaling in various vascular smooth muscle tissues.

## Conclusion

The peculiar vascular morphology of normal adrenal tissue and adrenal neoplasms may suggest a link between tissue specific milieu of adrenal glands/adrenal hormones/receptors and mechanism of pathogenesis of FMD, thus providing a direction of further investigation and management FMD in other anatomic sites.

## Data Availability

All data produced in the present work are contained in the manuscript.

## Author contribution

PZ reviewed the pathology and wrote the manuscript. DR and MR provided clinical information of cases. All authors read and approved the final manuscript.

